# Continuous glucose monitor–derived glucotypes and cardiovascular risk scores in individuals without diabetes

**DOI:** 10.64898/2026.02.25.26347035

**Authors:** Bahar Bakhshi, Honghuang Lin, Naznin Sultana, Elizabeth Healey, Havelah Queen, Sophie E. Claudel, Erika Minetti, Gary F. Mitchell, Joanne M. Murabito, Donald Lloyd-Jones, Devin W. Steenkamp, Matthew Nayor, Vanessa Xanthakis, Maura E. Walker, Nicole L. Spartano

## Abstract

**Introduction:** Dysglycemia is a well-established risk factor for cardiovascular disease (CVD); yet traditional glycemic traits, including fasting plasma glucose (FPG) and HbA1c, do not capture dynamic glucose fluctuations that may inform CVD risk. We cross-sectionally investigated the association of continuous glucose monitor (CGM)-derived metrics and 2-h post-prandial glucose (2-h PPG) with estimated 10-year CVD risk among individuals without diabetes.

**Methods:** We included 1,360 Framingham Heart Study participants (Third Generation, New Offspring Spouse, and Omni 2 cohorts at exam 4) without prevalent diabetes or CVD who had ≥3 days of CGM data and completed a mixed meal tolerance test (MMTT) with corresponding 2-PPG. We included 7 CGM summary metrics and defined data-driven glucotypes according to CGM measures of glycemic burden and variability. The 10-year CVD risk was estimated using the Predicting Risk of CVD EVENTs (PREVENT) base equations. We performed linear regression on standardized glycemic traits and glucotypes with log-transformed PREVENT risk scores and multinomial regression to relate standardized CGM metrics and 2-h PPG with PREVENT categories (low <5%[reference], borderline 5-<7.5%, intermediate/high ≥7.5%). All models were adjusted for FPG and body mass index (BMI).

**Results:** Among participants (55.9% women, 43.4% with prediabetes), mean age was 59.3 years, and mean BMI was 27.9 kg/m^2^. All CGM-derived metrics and 2-h PPG were positively associated with higher overall 10-year CVD risk (per 1 SD increase of each exposure variable, β range: 0.06-0.16, all p<0.001). A glucotype representing high glycemic burden and high glycemic variability was associated with higher overall 10-year CVD risk, compared with the glucotype representing low glycemic burden and low glycemic variability. Higher CGM-derived metrics and 2-h PPG were also associated with higher odds being in the intermediate/high CVD risk (OR range: 1.20-1.65, all p<0.001), adjusting for FPG and BMI.

**Conclusion:** Dynamic glycemic traits, including novel glucotypes that capture glycemic burden and variability, may provide novel insights into CVD risk prevention among individuals without T2D.

## INTRODUCTION

Dysglycemia is a continuous, yet phenotypically heterogeneous risk factor for cardiovascular disease (CVD), typically characterized for clinical purposes by static glucose thresholds.^1^ Even without progression to type 2 diabetes (T2D), impaired glycemic traits (i.e., fasting plasma glucose [FPG], HbA1c, or 2-h post-prandial glucose [PPG]) have been associated with adverse cardiometabolic outcomes, including atherosclerotic cardiovascular disease (ASCVD), heart failure (HF), chronic kidney disease (CKD), and all-cause mortality.^2,3^ Nonetheless, the existing T2D diagnostic framework oversimplifies the pathobiology underlying dysglycemia and its dynamic nature;^4^ impaired glycemic traits do not occur in isolation, but are rather broader manifestations of perturbations in glucose homeostasis and cardiometabolic regulation that may not be fully captured by traditional glycemic traits.^5-7^

Continuous glucose monitoring (CGM) can overcome these limitations by providing an opportunity for real-time assessment of glucose profiles, summarizing information on post-prandial glucose curves, temporal glucose dynamics, and overall glycemic burden and variability. While CGMs are now standard of care in diabetes management,^8^ large-scale CGM integration into population-based studies of individuals without diabetes has only recently started.^9-11^ Emerging studies among participants across the dysglycemia continuum have shown that CGM-derived summary measures and features of glucose curves (e.g., post-prandial glucose spikes) are correlated with cardiometabolic risk factors,^10^ and may offer incremental value in glycemic risk stratification beyond HbA1c alone.^9^ Harnessing dysglycemia heterogeneity, recent studies have also utilized CGMs’ rich analytical framework to identify glycemic subtypes (glucotypes) among those with and without T2D that represent phenotypic divergence in glycemic variability, hyper- and hypo-glycemia, and may reflect the underlying metabolic dysfunction, such as reduced insulin sensitivity as measured by low homeostatic model assessments of beta cell function.^12,13^ However, glucotypes identified among small studies of individuals without T2D have not been replicated in larger studies.^14,15^

Here, we present and characterize data-driven CGM-derived glucotypes, defined based on glycemic burden and variability, in a large dataset of individuals without T2D from the Framingham Heart Study (FHS). We also examine cross-sectional associations of CGM summary measures, as well as glucotypes, with the estimated 10-year CVD risk, using the Predicting Risk of cardiovascular disease EVENTs (PREVENT) equation.^16^ Our results generate complementary insights into dysglycemia-CVD relations that have not been characterized before.

## METHODS

### Study Sample

FHS Third Generation, New Offspring Spouse, and Omni 2 Cohorts attending their fourth examination cycle (2022–2025) were invited to undergo an oral mixed-meal tolerance test (MMTT) and wear a Dexcom G6 Pro CGM for up to 10 days (n=2718).^17^ Of those, we included participants who completed the blood draw pre- and post-MMTT and wore the CGM for ≥3 valid days following their visit. We excluded individuals with prior CVD events (up to 2022, n= 94), prevalent diabetes (defined as FPG ≥126 mg/dL or HbA1c ≥6.5%) or taking glucose-lowering medications (n=226), important covariates or PREVENT components missing or outside pre-defined ranges (n=206),^16^ resulting in a final sample size of 1360 (**Figure S1)**.

All FHS study protocols and procedures were conducted according to the guidelines of the Declaration of Helsinki and approved by the institutional review board for human research at Boston University. All participants provided written consent prior to participation in the study.

### Glycemic Traits

After a 10-hour overnight fast, participants were provided a liquid meal, Boost Plus challenge (MMTT, containing a total of 600 kcal, including 75 g carbohydrate, 21 g fat, 29 g protein), and were asked to consume within 5 minutes. Plasma samples were collected before and 2-h after the challenge (median time interval of 119 minutes) to measure FPG and 2-h PPG. Following the MMTT, participants were asked to wear a Dexcom G6 Pro monitor on their arm or abdomen for up to 10 days. Interstitial glucose values were measured every 5 minutes (max 288 readings/day) for the entire wear time (average wear time of 8.2 days). *CGM summary measures*. CGM data preprocessing was conducted according to steps detailed in **Figure S1** to define valid days of CGM wear. We used the interpreting blood glucose data (iglu) R package, a standardized, open-source package to calculate CGM summary measures.^18^ In our analysis, we included 7 CGM summary measures, based on previous literature, which can be broadly categorized into measures of overall glycemic burden and variability, including: mean glucose, J-index, percent of time spent above 140 mg/dL, coefficient of variation (CV), mean amplitude of glycemic excursions (MAGE), continuous overall net glycemic action over 1-hour (CONGA-1), mean of daily differences (MODD), and glycemic risk assessment diabetes equation (GRADE).^19,20^ The description of each summary measure is presented in **Table S1**.

### Ascertainment of Prior CVD Events

CVD is a composite outcome including coronary heart disease, congestive heart failure, stroke, and peripheral arterial disease. FHS outcomes were adjudicated via medical records that were reviewed by a panel of three experienced physicians using standard FHS criteria. A confirmed case of CVD was considered prevalent if the confirmed date of symptom onset occurred prior to 2022.^21,22^

### PREVENT Risk Estimates

We estimated the 10-year total CVD risk with PREVENT equations developed by the American Heart Association (AHA), which allows for sex-specific, race-free CVD risk calculation for US adults aged 30-79 years and previously described in detail.^16^ For our primary analysis, we used the base equations estimating the global CVD risk, incorporating traditional cardiometabolic risk factors within predetermined ranges, such as smoking status, systolic blood pressure (SBP, 90-200 mmHg), total cholesterol (TC, 130-320 mg/dL), high density lipoprotein (HDL, 20-100 mg/dL), antihypertensive medication use, statin use, diabetes status, in addition to estimated glomerular filtration rate (eGFR, ≥0 mL/min/1.73 m^2^).

### Statistical Analysis

#### CGM glycemic traits

Participant characteristics according to glycemic status were reported as mean (SD) for continuous variables and n (%) for categorical variables. Prediabetes was defined as either an HbA1c of 5.7-6.4% or FPG of 100-125 mg/dL. Age- and sex-adjusted pairwise correlation coefficients were used to evaluate the correlations between key glycemic traits.

To investigate the associations between glycemic traits and PREVENT risk estimates, we performed multivariable linear regression on standardized glycemic traits (mean 1 and SD of 0) and log-transformed PREVENT risk estimates, adjusting for FPG (model 1) and BMI (model 2). Because age, sex, cholesterol-lowering and anti-hypertensive medications are all components of PREVENT equations, we refrained from adjusting for these covariates to prevent over-adjustment, and subsequent bias toward the null.^23^

We categorized PREVENT scores to ACC/AHA risk categories, low risk <5.0%, borderline 5.0% to >7.5%, while collapsing the intermediate 7.5% to <20% and high risk ≥20% together to intermediate/high (≥7.5%).^24^ We performed multinomial regression on standardized glycemic traits (mean 1 and SD of 0) and PREVENT risk categories, with low risk as the referent, and adjusting for FPG and BMI.

#### Glucotypes

We defined glucotypes according to population-based median of glycemic burden (i.e., % time above 140 mg/dL) and variability (i.e., MAGE) metrics, on the basis of their robust linear associations with PREVENT risk estimates and their corresponding partial correlations being only moderate (see results). Participants were classified into four mutually exclusive groups based on placement below or above the median value of % time above 140 mg/dL or MAGE: 1. low burden, low variability, 2. low burden, high variability, 3. high burden, low variability, 4. high burden, high variability. We also included participant characteristics according to glucotype, reported as mean (SD) for continuous variables and n (%) for categorical variables (**Table S2**),

## RESULTS

Among participants without T2D (n=1,360), average age was 59.3 years (SD 8.4), 55.9% were female, 56.6% were normoglycemic, and average BMI was 27.9 (SD 4.4). Individuals with prediabetes (n=590) were older, had higher BMI and higher traditional glycemic traits, including FPG, HbA1c, 2-h PPG, as well as CGM summary measures, and more adverse cardiometabolic risk profiles, in comparison to individual with normoglycemia (n=790) (**Table 1**).

**Table 1.**
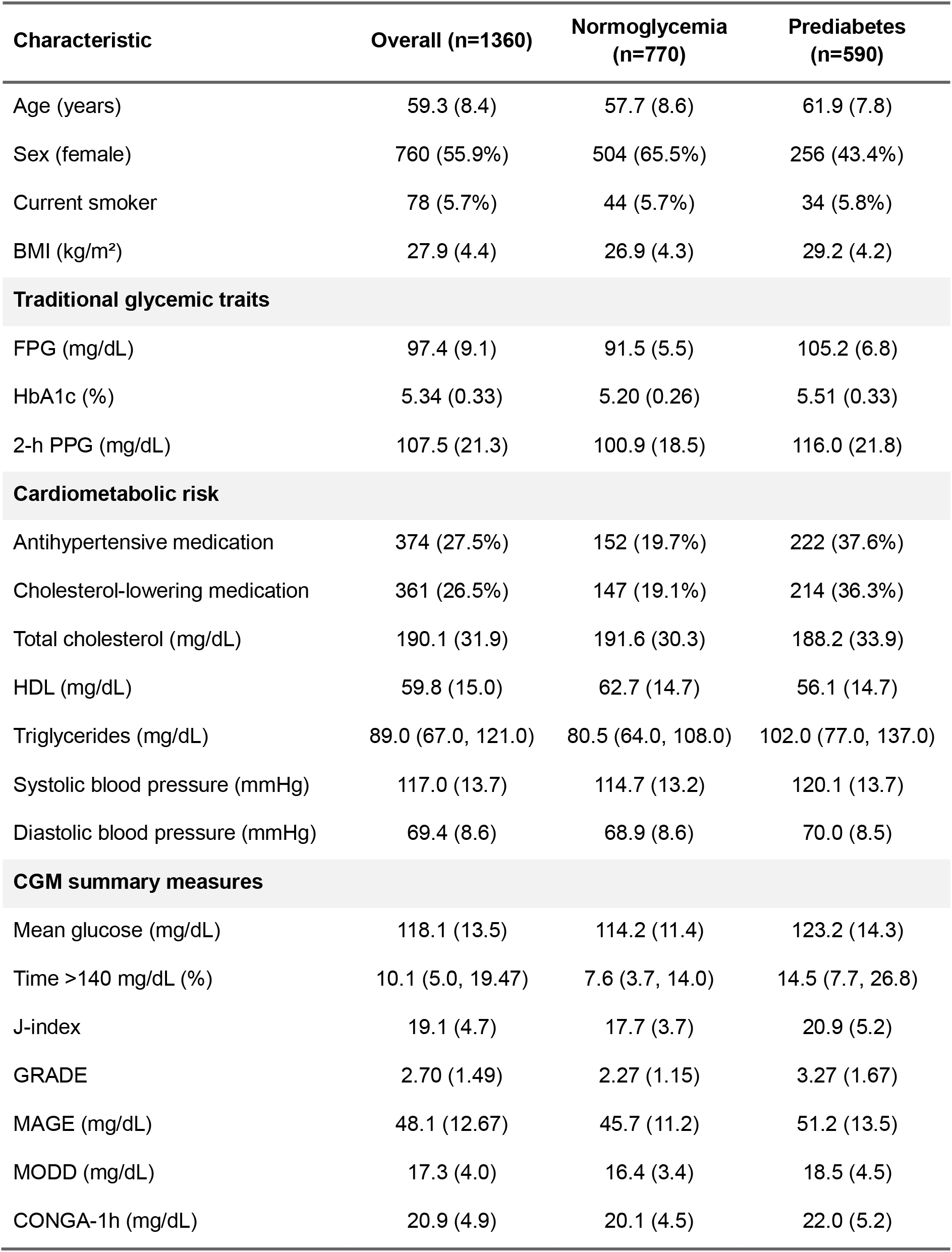
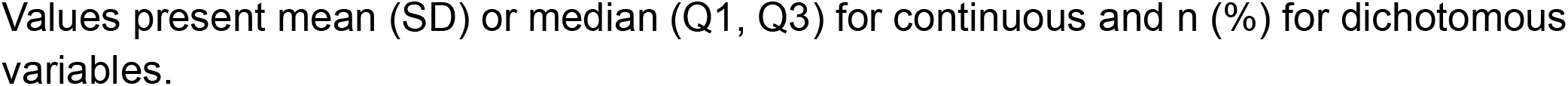
Participant characteristics according to glycemic status.

| Characteristic | Overall (n=1360) | Normoglycemia (n=770) | Prediabetes (n=590) |
| --- | --- | --- | --- |
| Age (years) | 59.3 (8.4) | 57.7 (8.6) | 61.9 (7.8) |
| Sex (female) | 760 (55.9%) | 504 (65.5%) | 256 (43.4%) |
| Current smoker | 78 (5.7%) | 44 (5.7%) | 34 (5.8%) |
| BMI (kg/m<sup>2</sup>) | 27.9 (4.4) | 26.9 (4.3) | 29.2 (4.2) |
| **Traditional glycemic traits** |  |  |  |
| FPG (mg/dL) | 97.4 (9.1) | 91.5 (5.5) | 105.2 (6.8) |
| HbA1c (%) | 5.34 (0.33) | 5.20 (0.26) | 5.51 (0.33) |
| 2-h PPG (mg/dL) | 107.5 (21.3) | 100.9 (18.5) | 116.0 (21.8) |
| **Cardiometabolic risk** |  |  |  |
| Antihypertensive medication | 374 (27.5%) | 152 (19.7%) | 222 (37.6%) |
| Cholesterol-lowering medication | 361 (26.5%) | 147 (19.1%) | 214 (36.3%) |
| Total cholesterol (mg/dL) | 190.1 (31.9) | 191.6 (30.3) | 188.2 (33.9) |
| HDL (mg/dL) | 59.8 (15.0) | 62.7 (14.7) | 56.1 (14.7) |
| Triglycerides (mg/dL) | 89.0 (67.0, 121.0) | 80.5 (64.0, 108.0) | 102.0 (77.0, 137.0) |
| Systolic blood pressure (mmHg) | 117.0 (13.7) | 114.7 (13.2) | 120.1 (13.7) |
| Diastolic blood pressure (mmHg) | 69.4 (8.6) | 68.9 (8.6) | 70.0 (8.5) |
| **CGM summary measures** |  |  |  |
| Mean glucose (mg/dL) | 118.1 (13.5) | 114.2 (11.4) | 123.2 (14.3) |
| Time >140 mg/dL (%) | 10.1 (5.0, 19.47) | 7.6 (3.7, 14.0) | 14.5 (7.7, 26.8) |
| J-index | 19.1 (4.7) | 17.7 (3.7) | 20.9 (5.2) |
| GRADE | 2.70 (1.49) | 2.27 (1.15) | 3.27 (1.67) |
| MAGE (mg/dL) | 48.1 (12.67) | 45.7 (11.2) | 51.2 (13.5) |
| MODD (mg/dL) | 17.3 (4.0) | 16.4 (3.4) | 18.5 (4.5) |
| CONGA-1h (mg/dL) | 20.9 (4.9) | 20.1 (4.5) | 22.0 (5.2) |

Values present mean (SD) or median (Q1, Q3) for continuous and n (%) for dichotomous variables.

### Pairwise Correlation of CGM metrics and Traditional Glycemic Traits

An age- and sex-adjusted heatmap of novel glycemic traits with traditional glycemic risk factors in the overall population is presented in **Figure 1**. CGM summary measures and 2-h PPG only weakly or moderately correlate with FPG and HbA1c among those without diabetes. While measures of overall glycemic burden (e.g., mean glucose and % time >140 mg/dL) and glycemic variability (e.g. MAGE and MODD) correlate strongly with measures within the same domain, measures of overall glycemic burden only partially correlate with measures of glycemic variability and vice versa.

**Figure 1.**
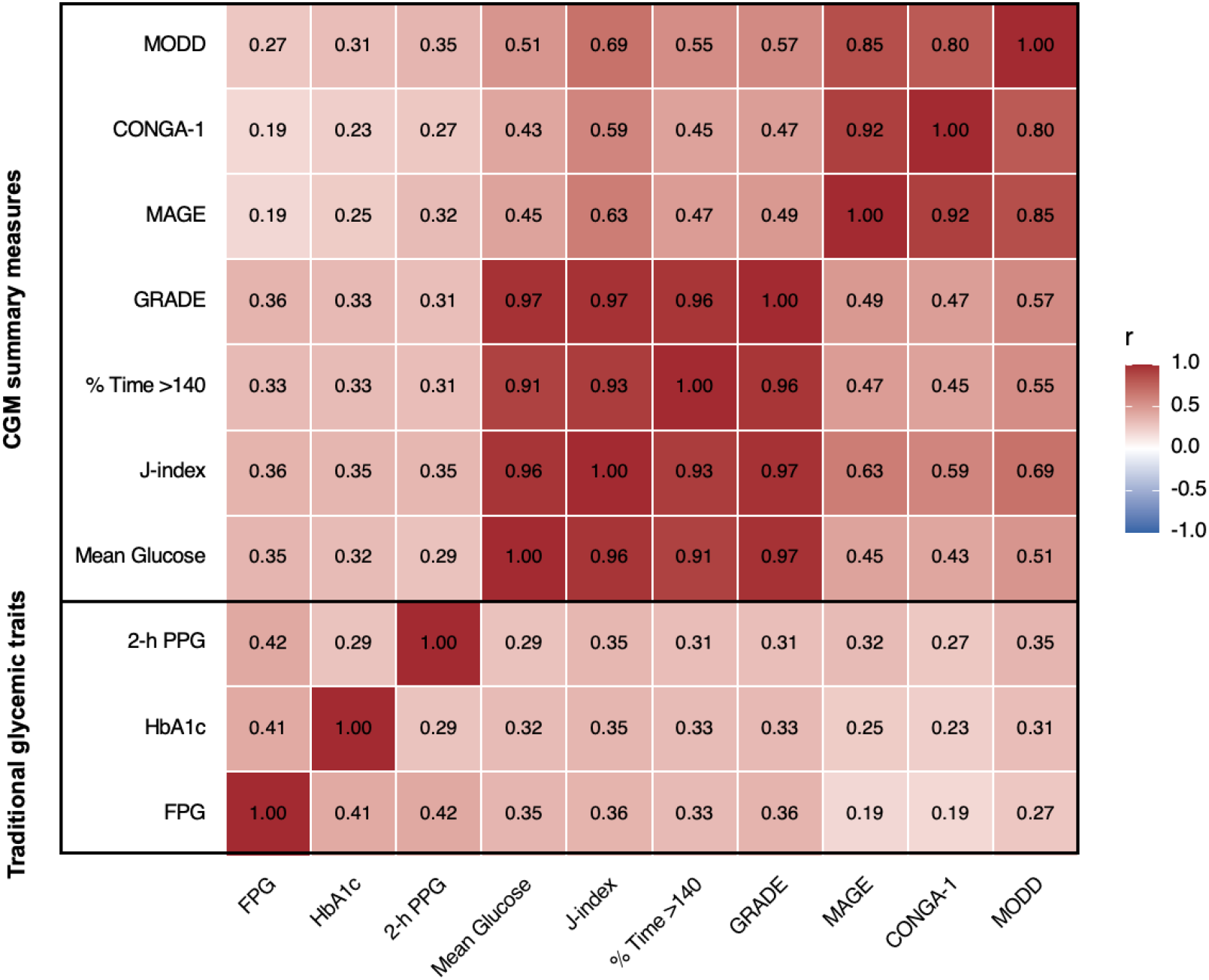
Age- and sex-adjusted correlations across traditional and CGM-derived glycemic metrics. Heatmap of Pearson correlation coefficients for FPG, HbA1c, 2-hour PPG, and continuous glucose monitoring (CGM) metrics of glycemic burden and variability. Each measure was first residualized for age and sex, and correlations were calculated using the adjusted values. Color intensity indicates the magnitude and direction of the correlation, and numeric values denote correlation coefficients (r). **Abbreviations:** FPG, fasting plasma glucose; HbA1c, hemoglobin A1c; 2-h PPG, 2-hour post– MMTT glucose; GRADE, Glycemic Risk Assessment Diabetes Equation; MAGE, mean amplitude of glycemic excursions; CONGA-1, continuous overall net glycemic action (1-hour); MODD, mean of daily differences.

### Glycemic Traits and 10-year Total CVD Risk Estimates

In the overall population, we observed significant associations of glycemic measures with log-transformed PREVENT–CVD (**Figure 2**), after adjusting for FPG. Higher 2-h PPG was associated with higher 10-year estimates for total CVD (β [SE] per 1 SD increase; 0.16 [0.12, 0.20]). Similarly, CGM measures were positively associated with higher 10-year estimates for total CVD (e.g., β [SE] per 1 SD increase; mean glucose: 0.07 [0.03, 0.11], log % time >140 mg/dL: 0.16 [0.06, 0.15], MAGE: 0.102 [0.06, 0.14], all p-values <0.001). The associations between glycemic measures and log-transformed PREVENT–CVD risk estimates persisted after further adjustment for BMI.

**Figure 2.**
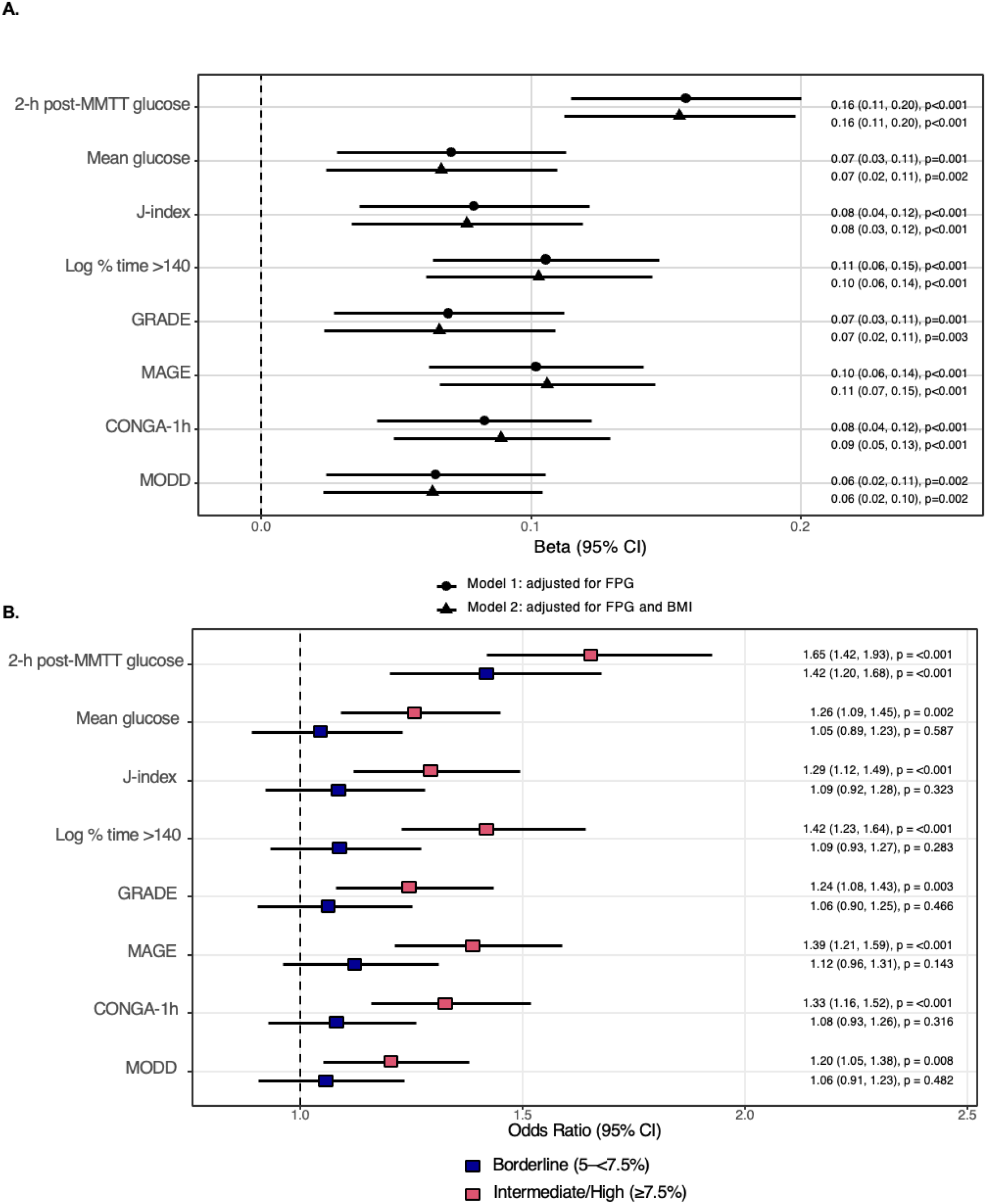
A. The associations of glycemic traits with 10-year total CVD risk estimates in the overall population (n=1360). Linear regression models were performed on standardized 2-h PPG and CGM summary measures with PREVENT scores, adjusting for FPG (model 1) and additional adjustment for BMI (model 2). Regression coefficient (betas) are presented on log scale and represent the change in log-transformed PREVENT scores per one unit increase in each exposure. **B. The associations of glycemic traits with odds of being in borderline (5-<7.5%, n=268) and intermediate/high PREVENT risk categories (**≥**7.5%, n=391), compared to low-risk category (<5%, n=701) as reference**. Multinomial regression models performed on standardized 2-h PPG and CGM summary measures and PREVENT scores, adjusting for FPG and BMI. **Abbreviations:** FPG, fasting plasma glucose; BMI, body mass index; 2-h PPG, 2-hour post– MMTT glucose; GRADE, Glycemic Risk Assessment Diabetes Equation; MAGE, mean amplitude of glycemic excursions; CONGA-1, continuous overall net glycemic action (1-hour); MODD, mean of daily differences.

In stratified models (**Figure S2**), we observed that most glycemic measures were positively associated with 10-year estimates for total CVD among individuals with normoglycemia and prediabetes, except for MAGE with associations only observed in those with prediabetes (e.g., normoglycemia; mean glucose: 0.07 [0.01, 0.14], log % time >140 mg/dL: 0.09 [0.03, 0.15], MAGE: 0.121 [0.06, 0.18] and prediabetes; mean glucose: 0.07 [0.01, 0.12], log % time >140 mg/dL: 0.11 [0.06, 0.17], MAGE: 0.09 [0.04, 0.14]).

Results from the multinomial regression indicated that after adjustment for FPG and BMI, higher 2-h PPG was associated with increased odds of being in the borderline PREVENT risk category (5-7.5%), as well as with increased odds of being in the intermediate/high PREVENT risk category (≥7.5%), compared with the low risk category as reference (OR [95% CI]: 1.65 (1.42, 1.93) and 1.45 (1.20, 1.68), respectively). While higher CGM summary measures were not associated with increased odds of being in the borderline PREVENT risk category, measures of glycemic burden (e.g., mean glucose) and variability (e.g., MAGE) were associated with higher odds of being in the intermediate/high PREVENT risk category (≥7.5%), compared with the low risk category as reference (OR [95% CI]: 1.26 [1.09, 1.45] and 1.39 [1.21, 1.43], for mean glucose and MAGE respectively).

### Glucotypes and 10-year Total CVD Risk Estimates

Across the four glucotypes, as traditional glycemic traits, including FPG, HbA1c, and 2-h PPG, gradually worsened, the prevalence of prediabetes increased: from 29.1% within the low burden, low variability group to 59.9% within the high burden, high variability group (**Figure 3**). Additionally, compared with the low burden, low variability group as reference, individuals in the other three glucotypes had higher predicted 10-year total CVD risk (β [SE] for low burden, high variability: 0.18 [0.06, 0.30]; high burden, low variability: 0.13 [0.01, 0.26]; high burden, low variability: 0.24 [0.15, 0.34]). Although the high burden, high variability group exhibited slightly higher PREVENT scores, the difference relative to other groups was not statistically significant (all pairwise comparisons p >0.05).

**Figure 3.**
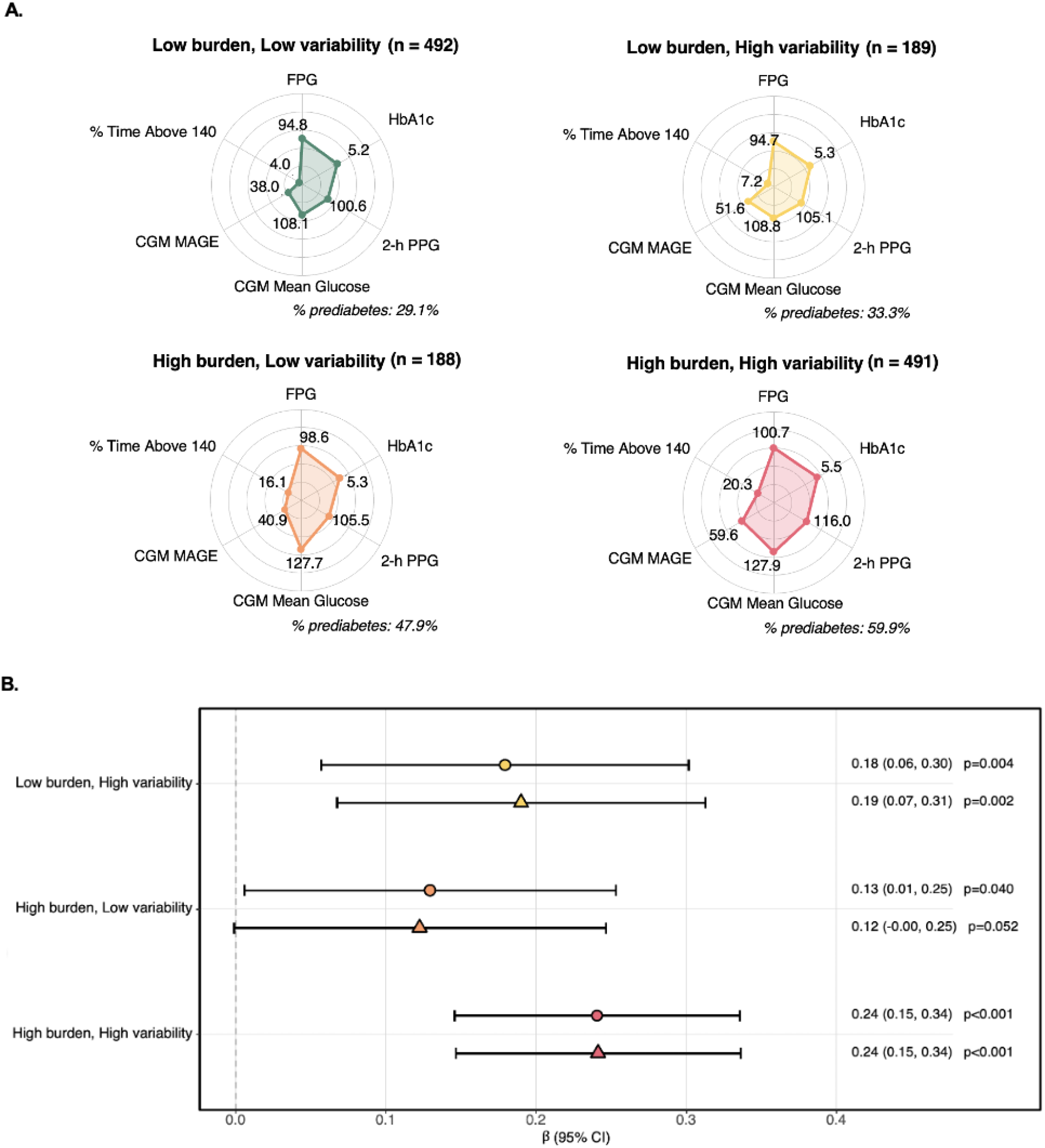
A. Glycemic profiles across glucotypes defined according to CGM glycemic burden and variability. Radar plots illustrating the mean values of FPG, HbA1c, 2-h PPG, mean glucose, MAGE, and median value of % time >140 across glycemic four burden– variability phenotypes. Prior to visualization, all features were min-max scaled (0-1 range), and prevalent prediabetes within each glucotype is presented. **B. The associations of glucotypes with PREVENT scores, compared to low burden, low variability as reference, in the overall population (n=1360)**. Linear regression models were adjusted for FPG (model 1) and additional adjustment for BMI (model 2). **Abbreviations:** FPG, fasting plasma glucose; HbA1c, Hemoglobin A1c; 2-h PPG, 2-hour post– MMTT glucose; MAGE, mean amplitude of glycemic excursions

## DISCUSSION

Despite ongoing preventive efforts to mitigate cardiometabolic risk, dysglycemia persists as a key contributor to CVD, independent from other frequently co-occurring metabolic abnormalities (e.g., central obesity, hypertension, and dyslipidemia) and progression to T2D.^2,3^ Traditional measures of glycemia lack granularity in capturing dynamic glucose fluctuations that occur as a result of daily physiologic perturbations. Here, we sought to investigate whether novel glycemic measures, reflective of continuous glucose dynamics, are cross-sectionally associated with estimated 10-year CVD risk among individuals without T2D, while considering relevant confounders. Our results indicate that among those without T2D, CGM summary measures and 2-h PPG are associated with higher 10-year estimated total CVD risk, and higher odds of being in the intermediate/high PREVENT risk categories, compared with the low-risk category. We also characterized CGM-derived glucotypes, according to metrics of glycemic burden and variability, demonstrating heterogeneity of glycemic profiles among a population without T2D. The glucotype representing the highest glycemic variability and burden was associated with higher estimated 10-year CVD risk, although differences relative to other groups were not statistically significant. Collectively, these findings support the concept that glycemic burden and variability can capture complementary but distinct dimensions of glycemia and may jointly identify individuals with progressively worsened cardiometabolic burden.

CGM has recently emerged as a tool to characterize temporal glucose trajectories of individuals without T2D, with an overarching aim of improving precision T2D prevention.^7^ Within Project 10k, a deeply phenotyped biobank of >7,000 non-diabetic participants, CGM-derived summary measures (e.g., mean glucose and MAGE) were consistently correlated with cardiometabolic risk factors, such as body composition and lipid profile in age- and sex- adjusted analysis, while some correlations were also observed with vascular- and liver-related outcomes.^10^ A smaller study, involving >300 individuals across the dysglycemia continuum, reported significant correlations of CGM-derived mean glucose and spike resolution (defined as the average time to absorb 50% of the glucose spike) with key contributors to cardiometabolic health, such as BMI, HbA1c, gut microbiome diversity.^9^ Another important study contributing to our understanding of the potential clinical value of CGM data in populations without diabetes is the A Estrada Glycation and Inflammation Study (AEGIS), the only cohort with both CGM data and longitudinal health outcomes. In this study, authors observed that higher % time >140 mg/dL, among individuals without T2D at baseline, was associated with incident T2D over 5 years of follow-up;^25^ and in longer follow-up, that CGM data may be more predictive of T2D development in comparison to HbA1c alone.^26^ Taken together, our results, in addition to prior literature, underscore the untapped clinical potential of utilizing CGMs for characterizing dynamic glycemic traits, identifying early glycemic dysregulation, and their relations with cardiometabolic outcomes.

Given that the existing CGM-metrics, such as those used in the current study, were predominantly developed in samples with diabetes where glucose excursions are often more pronounced,^27^ it is plausible that these measures may be limited in terms of capturing biologically meaningful variations in glucose levels among non-diabetic individuals. This limitation highlights the need for utilizing the entirety of CGM time series to create scalable, clinically interpretable CGM measures that address subclinical changes in glucose homeostasis to more advanced dysglycemic states. Although recent studies have used CGM data to define glucotypes within T2D samples,^12,13,28^ CGM-glucotyping efforts have been less investigated in samples without diabetes.^14,15^ A small study on 57 individuals (mostly without T2D) applied time series clustering algorithms to define clusters of “low”, “moderate”, “severe” variability clusters.^15^ While “severe” variability cluster was significantly correlated with insulin resistance, up to 39% of those with “severe” glycemic variability were missed by traditional glycemic traits, including FPG, HbA1c, and 2-h post-OGTT.^15^ However, replication efforts for defining these glucotypes in the larger Maastricht Study suggested the derived groups were not necessarily distinct phenotypes, but rather represented progression of glycemic burden (i.e., increasing mean glucose).^14^ Although the overarching premise of identifying early glucose dysregulation through CGM time series clustering is both compelling and warrants investigation, substantial knowledge gaps exist in establishing scalable and generalizable methods for defining actionable glucotypes in individuals without T2D.^7,14^ The glucotypes representing glycemic burden and variability, presented in our current work, were developed in a large cohort of individuals without T2D, but have not yet been replicated.

Our findings should be interpreted in light of some limitations. Although we included a robust sample size, FHS population predominantly consists of White participants with European ancestry who mainly reside in one geographical location, which may limit the external validity of our results. Additionally, we used PREVENT equations to estimate CVD risk, providing several methodological and clinical advantages, including but not limited to: 1) incorporating race-free, sex-specific risk estimation and improved risk prediction compared with the previously established equations (the pooled cohort equations [PCEs]),^16^ and 2) capturing a broader spectrum of cardiovascular-kidney-metabolic risk that may aid in identifying early cardiometabolic dysregulation.^29^ However, we have not measured urine albumin-to-creatinine ratio (uACR) in our samples, and therefore, cannot use PREVENT equations that additionally incorporate uACR, HbA1c, and social deprivation index to capture the entire cardiovascular-kidney-metabolic (CKM) spectrum. Due to a lack of longitudinal data, we cannot yet investigate the associations of novel glycemic traits, reflective of habitual glycemia, and long-term clinical CVD events among our participants. We additionally included glucose levels after an oral MMTT in our analysis, which may offer a more real-life, physiologically relevant representation of post-prandial metabolism, in comparison to an oral glucose tolerance test (OGTT).^30^ It is also worth mentioning that 2-h PPG, whether assessed by OGTT or MMTT, is a sensitive, well-validated marker of dysglycemia with well-established relations to T2D and CVD risk.^31,32^

In summary, we conclude that CGM metrics of glycemic variability and burden provide new insights into dysglycemia-CVD relations among those without T2D. The medical community has already begun to capitalize on CGM technology for improved diabetes management. Future work is warranted to determine if CGM may offer potential for shifting current clinical practice for T2D screening and prevention.

## Supporting information

All supplemental files

## Data Availability

All data produced in the present study are available upon reasonable request to the authors.

## Acknowledgements

We thank FHS participants for their continued support of the study.

## Funding

This investigation was supported by the Framingham Heart Study’s National Heart, Lung and Blood Institute contracts (N01-HC25195, HHSN268201500001I, 75N92019D00031) with additional support from NIDDK R01DK129305 and NHLBI R01HL156975. BB is supported by a Predoctoral Fellowship from the American Heart Association (26PRE1564902). EH is supported by NICHD T32HD040128. Dexcom also provided continuous glucose monitors at a discounted rate.

## Notes

### Competing Interest Statement

G.F.M. is the owner of Cardiovascular Engineering, Inc., a company that designs and manufactures devices that measure vascular stiffness. The company uses these devices in clinical trials that evaluate the effects of diseases and interventions on vascular stiffness. G.F.M. also serves as a consultant to and receives grants and honoraria from Novartis, Merck, Bayer, Servier, Philips, and deCODE genetics. G.F.M. is an inventor on pending patent applications that disclose methods for predicting various measures of biological age using pressure waveforms. DWS and NLS are consultants for Abbott Diabetes Care. DWS is also a clinical trial investigator for Abbott Diabetes Care.

### Funding Statement

This investigation was supported by the Framingham Heart Study National Heart, Lung and Blood Institute contracts (N01-HC25195, HHSN268201500001I, 75N92019D00031) with additional support from NIDDK R01DK129305 and NHLBI R01HL156975. BB is supported by a Predoctoral Fellowship from the American Heart Association (26PRE1564902). EH is supported by NICHD T32HD040128. Dexcom also provided continuous glucose monitors at a discounted rate.

### Author Declarations

Ethics committee/IRB of Boston Medical Center and Boston University Medical Campus gave ethical approval for this work.

