## Supplementary material for "Continuous glucose monitor–derived glucotypes and cardiovascular risk scores in individuals without diabetes": All supplemental files

**Figure S1**. Flowchart.

FHS Third Generation, New Offspring Spouse, and Omni 2 participants at their 4th examination cycle (n=2718)

Exclude those who:

– Were ineligible due to taking hydroxyurea (n=2)

– Did not take a CGM (n=306)

– Did not return a CGM (n=203)

– CGM device failure(n=56)

– No readable CGM data (n=6) or were worn for <1 day (n=44)

– Days with >20 % missing data (n=13) or flagged as outlier

(n=12) were considered invalid

– Completed <3 valid days of CGM data (n=54)

– Prior CVD events (n=94)

– Prevalent diabetes (n=226)

– Missing post-MMTT glucose or lipid profile(n=138)

– PREVENT components outside of the following ranges (n=206):

Age (30-79 yrs), total cholesterol (130-320 mg/dL), high density lipoprotein cholesterol (20-100 mg/dL), systolic blood pressure (90-200 mmHg), and body mass index (18.5-39.9 kg/m^2)

Final analytical sample (n=1360)

| **Table S1.** The definition of CGM summary measures. | |
| --- | --- |
| Mean glucose (mg/dL) | The average interstitial glucose levels during the entire wear time. |
| Time >140 mg/dL (%) | The proportion of monitored time in which interstitial glucose values are above 140 mg/dL.^1^ |
| J-index | An index combining mean and standard deviation (SD) of interstitial glucose to measure the quality of glycemic control:^2^  J-index = 0.001 × (mean + SD)^2^ |
| GRADE | A composite metric that assesses overall glycemic risk by applying a nonlinear weighting function to glucose values, emphasizing clinically important hypo- and hyperglycemic excursions.^3^ |
| MAGE (mg/dL) | A measure of within-day glycemic variability, averaging the magnitude of excursions between consecutive peaks (highest points) and nadirs (lowest points) during the wear time, while only counting the changes in glucose levels that are greater than 1 SD of the glucose for a 24-hour period.^4^ |
| MODD (mg/dL) | A measure of between-day glycemic variability, representing the average absolute differences in glucose levels at the same time point on consecutive days.^5^ |
| CONGA-1h (mg/dL) | Measure of within-day glycemic variability, representing the differences between consecutive glucose readings taken at 1-hour intervals across the wear time.^6^ |

**Figure S2. The associations of glycemic traits with 10-year total CVD risk estimates according to glycemic status.** Linear regression models were performed on standardized 2-h PPG and CGM summary measures and PREVENT scores, adjusting for FPG (model 1) and additional adjustment for BMI (model 2).

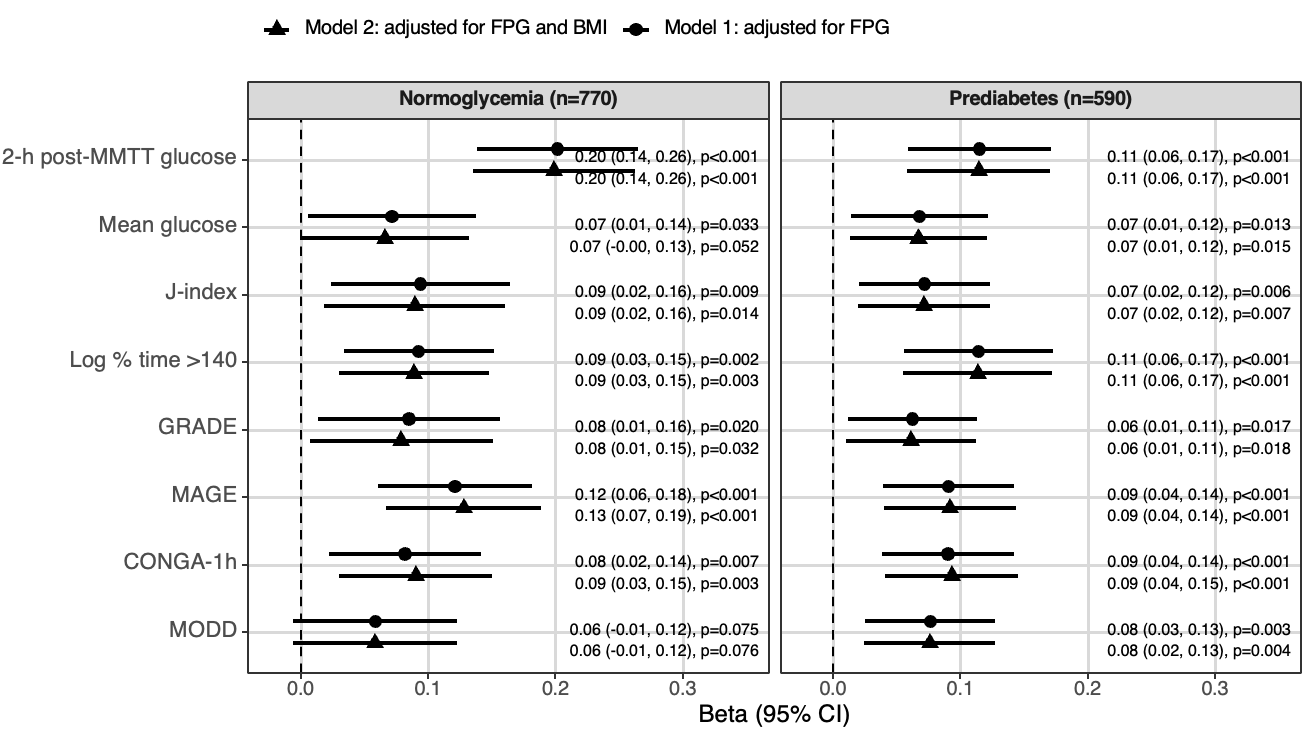

| **Table S2.** Participant characteristics according to glucotypes. | | | | | |
| --- | --- | --- | --- | --- | --- |
| **Characteristic** | **Overall (n=1360)** | **Low burden, low variability (n=492)** | **Low burden, high variability (n=189)** | **high burden, low variability (n=188)** | **high burden, high variability (n=491)** |
| Age (years) | 59.27 (8.43) | 57.63 (8.26) | 60.17 (8.37) | 58.48 (8.99) | 60.87 (8.06) |
| Sex (female) | 760 (55.9%) | 309 (62.8%) | 127 (67.2%) | 82 (43.6%) | 242 (49.3%) |
| Current smoker | 78 (5.7%) | 32 (6.5%) | 17 (9.0%) | 6 (3.2%) | 23 (4.7%) |
| BMI (kg/m²) | 27.91 (4.43) | 27.62 (4.24) | 26.47 (4.43) | 28.90 (4.19) | 28.37 (4.55) |
| **Traditional glycemic traits** | | | | | |
| FPG (mg/dL) | 97.41 (9.12) | 94.76 (8.14) | 94.69 (8.40) | 98.63 (7.47) | 100.66 (9.73) |
| HbA1c (%) | 5.34 (0.33) | 5.24 (0.29) | 5.27 (0.29) | 5.33 (0.29) | 5.47 (0.35) |
| 2-h PPG (mg/dL) | 107.46 (21.33) | 100.56 (17.37) | 105.10 (18.67) | 105.50 (16.28) | 116.03 (24.48) |
| Prediabetes status | 590 (43.4%) | 143 (29.1%) | 63 (33.3%) | 90 (47.9%) | 294 (59.9%) |
| **Cardiometabolic risk** | | | | | |
| Antihypertensive medication, | 374 (27.5%) | 116 (23.6%) | 35 (18.5%) | 55 (29.3%) | 168 (34.2%) |
| Lipid-lowering medication | 361 (26.5%) | 103 (20.9%) | 40 (21.2%) | 47 (25.0%) | 171 (34.8%) |
| Total cholesterol (mg/dL) | 190.13 (31.93) | 190.76 (30.86) | 194.84 (31.27) | 188.96 (34.76) | 188.14 (32.00) |
| HDL (mg/dL) | 59.84 (15.02) | 62.25 (14.22) | 64.74 (15.12) | 54.36 (13.78) | 57.63 (15.22) |
| Triglycerides (mg/dL) | 89.00 (67.00, 121.00) | 82.00 (63.00, 112.50) | 80.00 (63.00, 105.00) | 102.50 (71.00, 135.50) | 95.00 (73.00, 129.00) |
| Systolic blood pressure (mmHg) | 117.04 (13.66) | 115.00 (12.61) | 114.33 (13.48) | 118.57 (14.84) | 119.55 (13.78) |
| Diastolic blood pressure (mmHg) | 69.38 (8.57) | 69.17 (7.95) | 67.68 (8.63) | 71.58 (9.46) | 69.39 (8.63) |
| **CGM glycemic traits** | | | | | |
| Mean glucose (mg/dL) | 118.09 (13.50) | 108.12 (7.61) | 108.80 (5.59) | 127.75 (8.49) | 127.95 (11.92) |
| Time >140 mg/dL (%) | 10.12 (5.03, 19.37) | 4.03 (1.97, 6.11) | 7.18 (5.57, 8.70) | 16.14 (12.50, 23.80) | 20.31 (14.35, 31.30) |
| J-index | 19.05 (4.68) | 15.32 (1.97) | 16.48 (1.37) | 21.02 (2.61) | 23.03 (4.42) |
| GRADE | 2.70 (1.49) | 1.59 (0.56) | 1.80 (0.39) | 3.63 (1.09) | 3.82 (1.47) |
| MAGE (mg/dL) | 48.10 (12.57) | 38.01 (5.05) | 51.57 (5.48) | 40.87 (3.83) | 59.64 (11.61) |
| MODD (mg/dL) | 17.32 (4.04) | 14.34 (2.01) | 17.46 (2.08) | 15.79 (1.90) | 20.83 (4.00) |
| CONGA-1h (mg/dL) | 20.91 (4.90) | 17.09 (2.55) | 22.34 (2.63) | 18.28 (2.18) | 25.20 (4.36) |

Values present mean (SD) or median (Q1, Q3) for continuous and n (%) for dichotomous variables.
